# Leishmaniasis in migrants crossing Central America and México: An emerging epidemiological threat

**DOI:** 10.64898/2026.01.20.26344417

**Authors:** Jaime Zamora-Chimal, Mario A Rodríguez-Pérez, Isabel C. Cañeda-Guzmán, Lihua Wei, Adriana Ruiz-Remigio, Gabriela Meneses-Ruiz, José P. García-Ruíz, Norma Salaiza, Javier Juárez-Gabriel, Jesús A. Aguilar-Durán, Xianwu Guo, Uriel Hernández-Salinas, Edith G. Garrido-Lozada, Sarah A. Hamer, Gabriel L. Hamer, Ingeborg Becker, Nadia A. Fernández Santos

## Abstract

**Background:** Irregular migration across Central America and Mexico has increased in recent years, exposing migrants to various health risks, including vector-borne diseases such as leishmaniasis. The migration corridor from South America, through the Darién Gap, to northern Mexico traverses multiple leishmaniasis-endemic regions with active *Leishmania* spp. transmission, where environmental exposure during migrant’s transit may increase the risk of infection.

**Methodology / Principal Findings:** From 2023 to 2025, clinical and epidemiological data were collected from 365 migrants transiting through Mexico, including a subset of individuals with clinical suspicion of cutaneous leishmaniasis (CL). Migrants were evaluated at two shelters in northern Mexico and at the Centro de Medicina Tropical (CMT) of the Universidad Nacional Autonoma de Mexico (UNAM). Diagnostic testing included serology and polymerase chain reaction (PCR) for species identification. Sociodemographic data, country of origin, and migration routes were documented. A complementary literature review was conducted to map the geographic distribution of sand fly vectors along migratory pathways.

A total of 26 leishmaniasis clinical cases were identified, namely a 7.1% prevalence (95%-CI = 4.6-9.9%). Most individuals originated from Venezuela (n = 12), Honduras (n = 3), and Mexico (n = 3), with additional cases from Haiti and the Czech Republic. Six participants did not disclose their country of origin but reported traveling from South America. Polymerase chain reaction confirmed active infections with *Leishmania (Viannia) panamensis* (7 cases) and *Leishmania (Viannia) braziliensis* (1 case), both detected among Venezuelan migrants. Additional PCR-positive cases were classified within the *Viannia* subgenus.

**Conclusions/Significance:** These findings underscore the urgent need for tailored surveillance strategies and integrated cross-border healthcare interventions to address migrant populations, who may also contribute to the introduction of *Leishmania* species into Mexico, including those belonging to the *Viannia* subgenus. The presence of competent sand fly vectors along migration routes heightens the risk of transmission to both migrants and local resident communities.

**Author Summary:** Leishmaniasis is a neglected tropical disease transmitted by sand flies which remains a major public health concern in many parts of Mexico, Central, and South America. In recent years, irregular migration from South America to northern Mexico has increased, exposing migrants to endemic disease risks. Here, clinical and epidemiological data from migrants transiting Mexico between 2023 and 2025 were examined, and 26 cases of cutaneous leishmaniasis were identified. Species-level identification confirmed infections with *L.* (*V*.) *panamensis* and *L.* (*V*.) *braziliensis*, underscoring the importance of precise diagnosis to help guide treatment and clinical follow-up of migrants. The presence of competent sand fly vectors along migration routes further highlights the vulnerability of the migrant population. These findings emphasize the urgent need for active surveillance and integrated healthcare strategies to protect both migrant and local populations, while also considering social and ecological factors. Addressing leishmaniasis in an integrated context requires coordinated medical and educational interventions to prevent transmission, reduce disease burden, and support vulnerable communities.

## Introduction

Migration in Latin America has intensified in the last 20 years driven mainly by war, violence, armed conflict, poverty, political instability, and environmental change (1). The countries contributing the highest number of migrants include Venezuela, Honduras, Guatemala, Ecuador, Haiti, Cuba, Colombia, and El Salvador, with many individuals crossing Mexico in route to the United States. The principal route of migrants from South American passes through the Darién Gap between Colombia and Panama, a region with high transmission of leishmaniasis (1,2).

Leishmaniasis is a parasitic disease caused by a protozoan of the genus *Leishmania* and transmitted by the bite of female sand flies (Diptera: Psychodidae: Phlebotominae) (3). Globally, 21 *Leishmania* species are known to infect humans (4). In Latin America, infections are mainly caused by species of the subgenus *Leishmania* (*L. mexicana*, *L. amazonensis,* and *L. infantum*), and the subgenus *Viannia* [*L.* (*V*.) *braziliensis*, *L.* (*V*.) *guyanensis*, *L.* (*V*.) *panamensis,* and *L.* (*V*.) *peruviana*] (4,5). All of these species can cause cutaneous leishmaniasis (CL) (4), while *L.* (*V*.) *braziliensis*, *L.* (*V*.) *panamensis,* and *L.* (*V*.) *guyanensis* may also cause mucocutaneous leishmaniasis (6).

Currently, over 1,060 sand fly species have been described worldwide of which 556 occur in the Americas; 48.7 % of these are found in Brazil. If added to sand fly species from countries of Amazon areas such as Bolivia, Colombia, Ecuador, French Guiana, and Venezuela, they account for 95.2%. In Panama and Mexico, 71% and 75% of the known species, respectively, have been recorded (7). Some species have wide distributions such as *Psychodopygus panamensis*, *Lutzomyia gomezi,* and *Nyssomyia ylephiletor,* which are considered vectors of *L.* (*V*.) *braziliensis* and *L.* (*V*.) *panamensis* (8–12). The present study assesses leishmaniasis cases among migrants at two destination sites in Mexico from 2022 to 2025. In addition, a mini-review of literature on sand fly vector species involved in the transmission of *Leishmania* spp. relevant to migrant populations is presented.

## Materials and methods

### The study area and migrant populations

This study comprised two groups. The first one included 339 migrants evaluated at two shelters in Reynosa, Tamaulipas, Mexico (“Casa del Migrante Nuestra Señora de Guadalupe” and “Senda de Vida”) from July 2022 to October 2023. The second group consisted of 26 individuals with cutaneous leishmaniasis (CL), who were referred to the Centro de Medicina Tropical (CMT) in the Facultad de Medicina of the Universidad Nacional Autonoma de Mexico (UNAM) during 2024.

Participants were eligible if they met the sole inclusion criterion of being adult migrants aged 18 years and older. Each participant completed a structured questionnaire for collecting his/her data, clinical history, and their knowledge about vector-borne diseases. The questionnaire comprised 46 variables, including demographic data such as full name, date of birth, country of origin, gender, height, weight, age, duration of stay at the shelter, and countries and time periods spent traveling from their point of origin to destination in Reynosa. In addition, the questionnaire assessed knowledge about arthropod vectors, vector-borne diseases, and the preventive measures used to avoid infection. The study protocol was approved by the Ethics Committee of the Escuela Nacional de Medicina y Homeopatia of the Instituto Politécnico Nacional (Reference number: IRB No. CBE/006/2020). All adult participants provided written informed consent prior to enrollment.

### Samples for leishmaniasis diagnosis

The diagnosis of cutaneous leishmaniasis was based on epidemiological and clinical history, combined with the detection of the parasite through skin scraping, culture or molecular identification of parasite DNA. *Leishmania* species were identified by polymerase chain reaction (PCR) followed by DNA sequencing. Diagnostic approaches varied across study populations: in migrants examined at the CMT in Mexico city, molecular methods were used to detect *Leishmania* DNA, whereas migrants assessed at shelters in Reynosa underwent only serological testing with ELISA for anti-*Leishmania* antibodies.

For serological testing, venous blood was collected using a 21G needle (BD Vacutainer) and serum separator tubes (6.0 mL, BD Vacutainer). Samples were maintained on ice, centrifuged at 2,000 rpm for 15 minutes at -5°C and serum aliquots were stored in 1.5 mL Eppendorf tubes at -70°C until analysis.

The in-house ELISA was performed using *Leishmania mexicana* promastigote lysate as antigen on 96-well plates. After blocking with casein, patient sera (1:100) were added in triplicate, followed by incubation with an HRP-conjugated anti-human antibody mixture (IgA, IgG, IgM, κ, and λ). Detection was carried out with TMB substrate, and absorbance was read at 450 nm. The cut-off was defined as the negative control mean plus three standard deviations. Results were classified as positive (≥10% above cut-off), negative (≤10% below cut-off), or indeterminate (within ±10% of cut-off).

### DNA extraction, PCR, and sequencing

DNA was extracted from parasite cultures obtained from cutaneous ulcers samples using the DNEasy Blood & Tissue Kit (QIAGEN Inc., Hilden, Germany), following manufacturer’s instructions. A ∼400 bp fragment of the ITS2 region was amplified with primers LGITSF2 (GCATGCCATATTCTCAGTGTC) and LGITSR2 (GGCCAACGCGAAGTTGAATTC), as previously described for *Leishmania* species identification (13). The PCR was performed using the AmpliTaq Gold™ DNA Polymerase LD kit (Applied Biosystems, Foster City, CA, USA). Amplicons were purified and sequenced bidirectionally using the BigDye™ Terminator v3.1 Cycle Sequencing Kit on a 3500 Genetic Analyzer (Applied Biosystems). Sequence analysis of PCR positive products was conducted at the Instituto de Diagnostico y Referencia Epidemiologicos of the Secretaria de Salud in Mexico city.

### Literature-based compilation of vector and parasite distribution

To contextualize the epidemiological relevance of *Leishmania* species identified in migrant patients, a literature mini-review was conducted. Data on phlebotomine sand flies associated with *Leishmania* transmission were compiled for endemic areas along migratory routes, including Panama (Darién Gap), Guatemala, Belize, and southern Mexico (Chiapas and Quintana Roo).

Peer-reviewed scientific articles and official public health reports were reviewed to identify vector species implicated in the transmission of *L.* (*V*.) *panamensis* and *L.* (*V*.) *braziliensis* in each country (8, 9, 14–18). The data were then used to map vector distributions and depict the potential exposure to *Leishmania* vectors along different regions of the migratory routes. The map was generated using QGIS v3.28.2. Free administrative-boundary GIS data for Mexico were obtained from the Instituto Nacional de Estadística y Geografía (INEGI; https://www.inegi.org.mx/app/mapas/). Satellite imagery and base maps available in QGIS, as well as Google Maps (https://www.google.com/maps) were used as visual references.

### Statistical Analysis

Descriptive statistical analyses were performed using Microsoft Excel of Windows to calculate prevalence of infection (number of positive individuals/total examined × 100), percentage of migrants per nationality and gender (number of individuals per country/total migrant population × 100), median of age, and range. The proportion of individuals harboring anti*-Lehismania* antibodies was calculated as the number of positive individuals divided by the total number examined and expressed as a percentage. The method of Armitage and Berry (1994) was used to estimate the 95% exact confidence intervals (CI) surrounding the point seroprevalence (19).

## Results

A total of 339 migrants were surveyed from July 2022 to October 2023 at the “Casa del Migrante Nuestra Señora de Guadalupe” and “Senda de Vida” shelters in Reynosa. Participants originated primarily from Honduras (25%), Haiti (20%), Mexico (19%), and Venezuela (17%), with a smaller proportion from El Salvador (8%), Colombia (4%), Guatemala (4%), Cuba (1%), Nicaragua (1%), Ecuador (0.6%), Dominican Republic (0.2%), and Brazil (0.2%). The median age of migrants was of 31 years old (range: 25 to 49 years old), with females comprising 63% of the total sample. However, among Venezuelan migrants, the proportion of males was higher than that of females (Figure 1).

**Figure 1.**
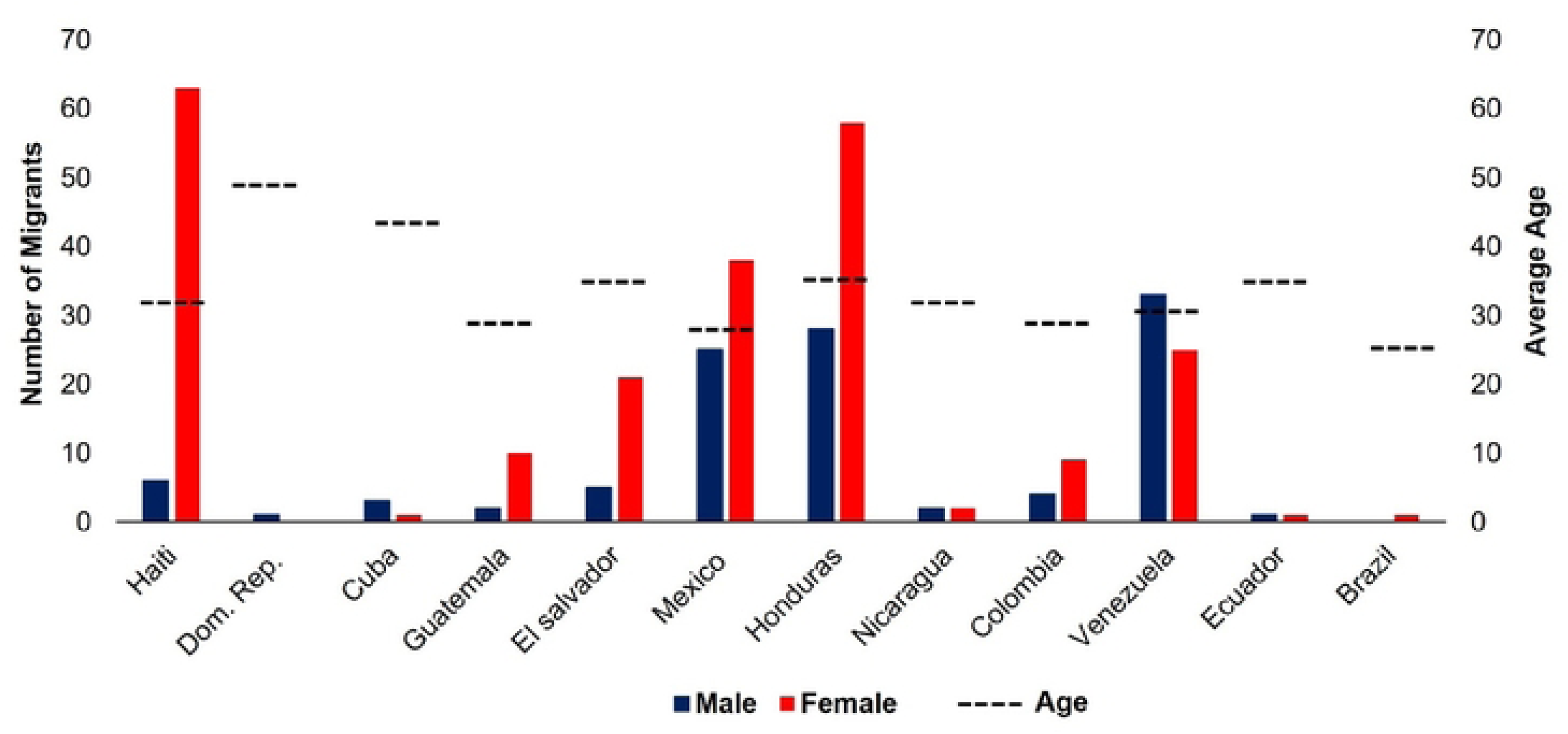
Country of origin and age group of the migrants (n=339) examined from 2022 to 2023 at the “Casa del Migrante” in Reynosa, México.

### Perceptions of vector exposure, health risk awareness, and social vulnerability among migrants during transit

A total of 339 migrants responded to questions regarding exposure to arthropod vectors, awareness of vector-borne diseases, and experiences of social vulnerability during transit (Figure 2). Overall, 65% reported contact with mosquitoes during their journey, and 71% were aware of mosquito-borne diseases. In contrast, only 36% recognized kissing bugs, whereas 86% recognized ticks. Regarding access to healthcare, 60% reported having knowledge of health services during transit. Notably, 35% of migrants reported experiencing discriminatory events during their journey.

**Figure 2.**
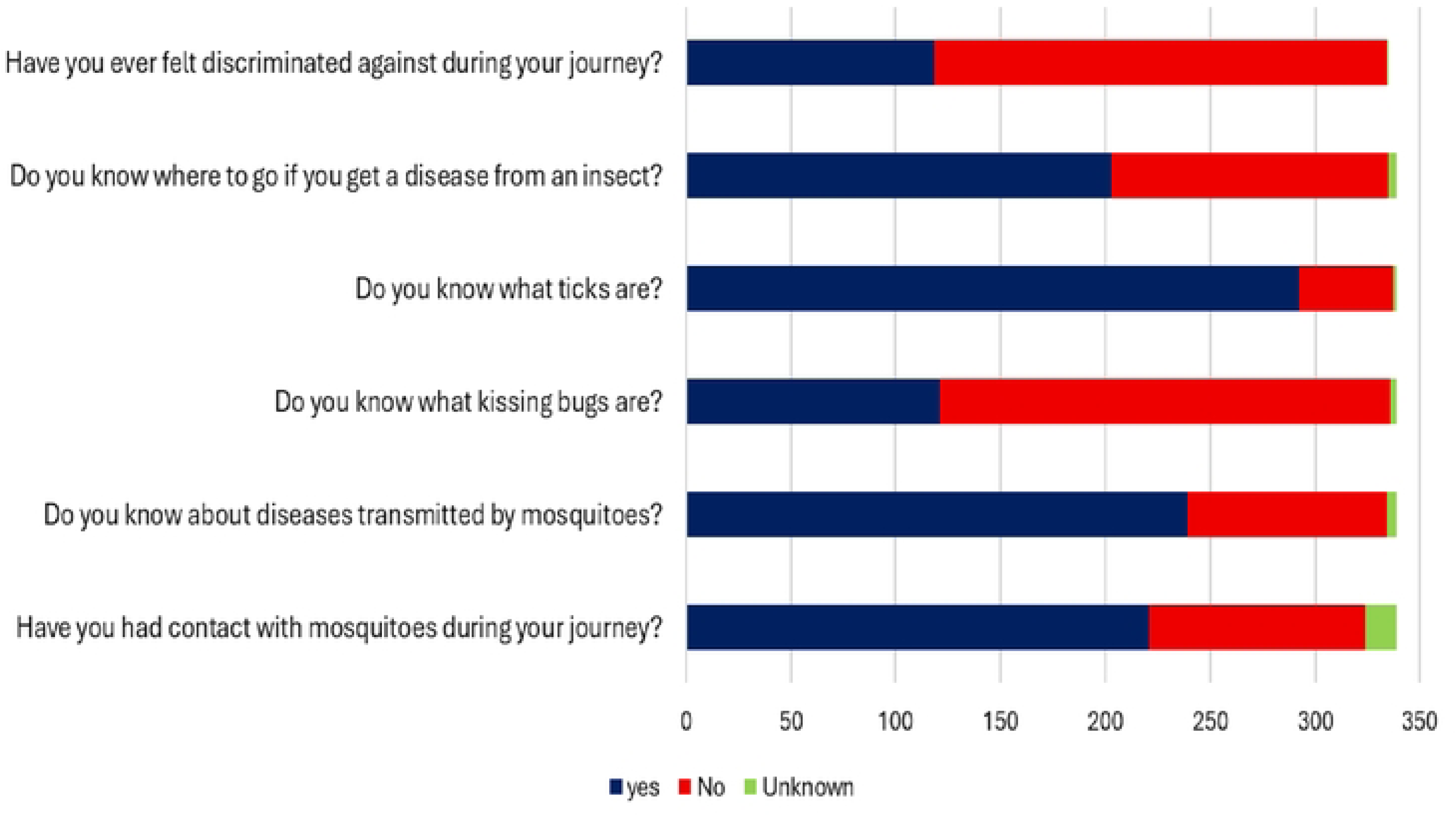
Number of migrant respondents and their responses regarding arthropod vectors and the diseases they transmit, social vulnerability (specifically discrimination), and vector exposure during transit. A total of 339 respondents from 2022 to 2023 participated at “Casa del Migrante Nuestra Señora de Guadalupe” and “Senda de Vida” in Reynosa, México.

### Migrants diagnosed with leishmaniasis during transit through Mexico

Overall, 26 clinical cases of leishmaniasis were identified among 365 migrants examined between 2023 and 2025, corresponding to a prevalence of 7.1% (95%-CI = 4.6-9.9%; Table 1). Of these, 23 clinical cases involved migrants transiting through Mexico in route to the United States. Two additional cases recorded in Mexico city and one from the Czech Republic in 2024 were infected during their stays in Peru and Costa Rica. Twelve patients were positive for *Leishmania* by ELISA, representing a seroprevalence of 3.6% (95%-CI = 1.8%-5.7%) of a total of 339 migrants examined in Reynosa shelters. These cases corresponded to individuals from Venezuela, Honduras, Haiti, and Mexico. The remaining 14 of the 26 migrants (53%; 95%-CI = 35%-71%) were diagnosed using different methods, including parasite isolation for DNA extraction, PCR, and sequencing. Molecular analysis confirmed infections with *Leishmania* (*V.*) *braziliensis* and *L.* (*V*.) *panamensis*. Half of the cases were positive for *L.* (*V.*) *panamensis* (n= 7), predominantly among Venezuelan migrants; 43% were classified within the *Viannia* complex without species-level resolution; and 7% were infected as *L.* (*V.*) *braziliensis* (Table 1). The remaining leishmaniasis cases, only diagnosed by ELISA, could not be attributed to a given species.

### Migratory route to the United States and contact with sand flies

Most Venezuelan and Caribbean migrants reported a migratory route crossing seven countries: Panama, Costa Rica, Nicaragua, El Salvador, Honduras, Guatemala, and Mexico. The Darién Gap in Panama was consistently identified as the most difficult and dangerous section of the journey. This rainforest region harbors a high biodiversity, where migrants are potentially exposed to multiple biting insects, including sand fly vectors for *L.* (*V.*) *panamensis* and *L.* (*V.*) *braziliensis* (Figure 3).

**Figure 3.**
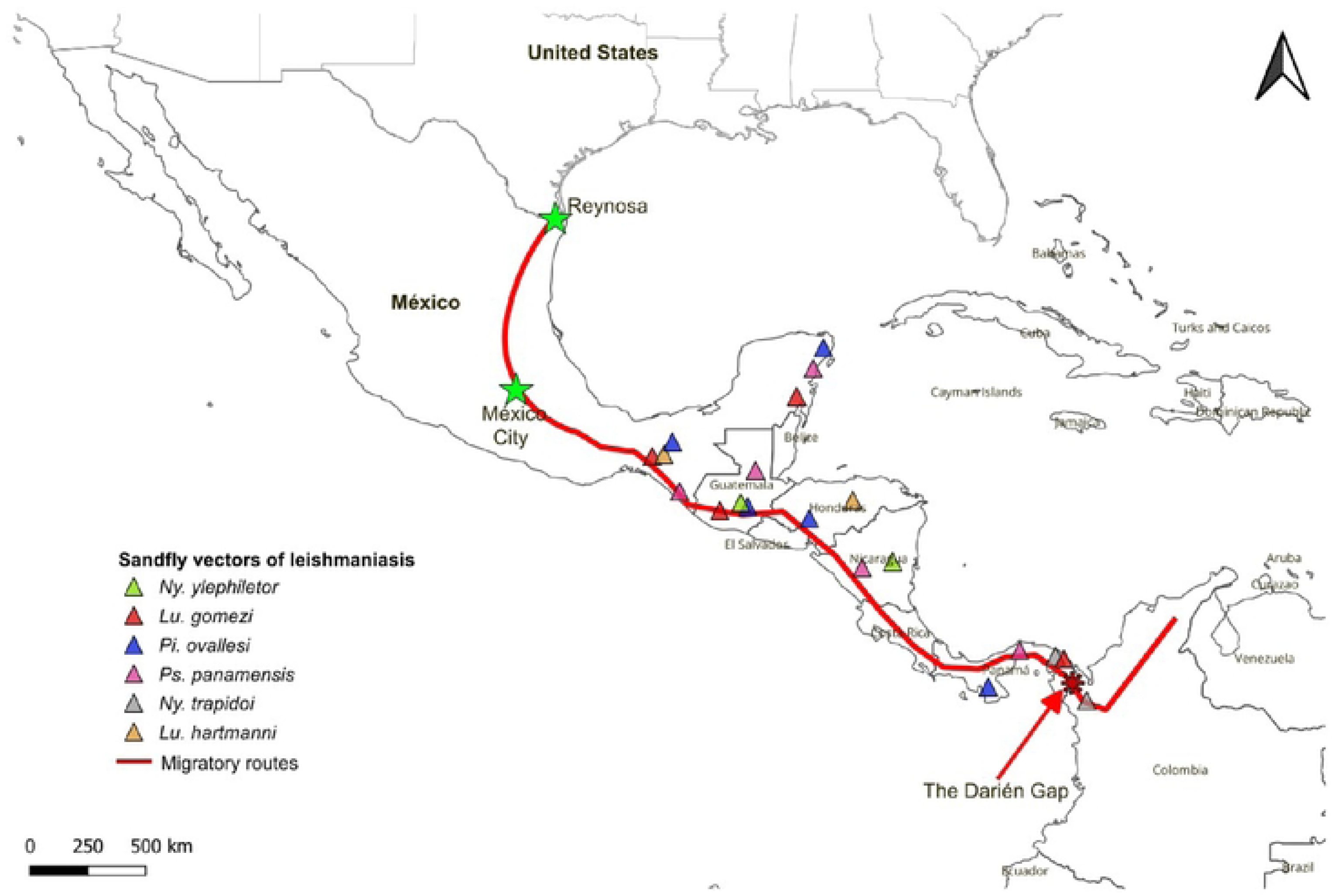
Migratory route from Venezuela to the U.S. border and potential exposure to *Leishmania* vectors. Triangular markers indicate the distribution of the six anthropophilic sand fly species that transmit *L.* (*V.*) *braziliensis* and *L.* (*V.*) *panamensis*: *▴ Nyssomyia trapidoi, ▴ Ny. ylephiletor, ▴ Psychodopygus panamensis, ▴ Pintomyia ovallesi, ▴ Lutzomyia Hartmanni,* and *▴ Lu. gomezi.* The red line indicates the possible migratory routes followed by migrants. It is worth noting that the sampling sites were located at the “Casa del Migrante in Reynosa” in northestern Mexico, and in Mexico City (marked with green stars). The red asterisk shows the Darién Gap.

### Species of sand flies that transmit Leishmania (V.) braziliensis and L. (V.) panamensis

*Lehismania* (*V.*) *panamensis* is the predominant species in Panama and can be transmitted by at least by five sand fly species. In contrast, *L.* (*V.*) *braziliensis* is the predominant species in Guatemala and can be transmitted by four sand fly species (Table 2). However, only few studies for *L.* (*V.*) *braziliensis* in Belize and similarly limited data are available for the border states of Mexico (Chiapas y Quintana Roo). Interestingly, *Lu. gomezi, Ny. ylephiletor*, *Ps. panamensis, Pi. ovallesi*, and *Lu. hartmanni* show a wider distribution across Central America extending into Mexico (Table 2).

## Discussion

This is the first comprehensive cross-sectional study of leishmaniasis of migrants in transit route from the America southern hemisphere to northern Mexico, crossing Central America. During the 2023–2025 period of this study, a higher proportion of migrant females was observed. This is concordant with data from the International Organization for Migration (20), which reported that during 2020, the gender distribution among individuals emigrating from Latin American and Caribbean countries to northern latitudes showed a predominance of females (20). The overrepresentation of females in our sample examined may be explained by several contextual social factors, including domestic violence, femicide, gang territorial control, and the weakening of state institutions reported in many South and Central American and Caribbean countries. These factors have contributed to an increase in forced female migration, often accompanied by dependent minors (21).

The median age of migrants by country in this study also falls within the 25-to-49-years old age group, suggesting that most are of working age and likely seeking better economic opportunities. Overall, the working-age range among migrants generally spans from 18 to 64 years (22).

Regarding knowledge about arthropod vector and the diseases they transmit, and social vulnerability, the 66.7% of the respondents reported frequent insect bites, and 39.5% indicated that they knew where to seek medical attention if they became ill. On average, migrants reported spending about one week in the Darién region, a duration that has remained consistent over the years, typically ranging from 5 to 15 days (23). After crossing the Darién, migrants continued their journey northward to the United States in seek of asylum. Knowledge about disease arthropod vectors varied among respondents: while 70.5% recognized mosquito-borne diseases, only 35.6% were able to identify triatomine bugs (“kissing bugs”), in contrast to 86% who were able to recognized ticks. In addition, 34.8% of migrants reported experiencing discrimination. This not only affects psychosocial well-being but may also have direct health implications, including potential immunological effects (24, 25).

It is worth noting that of 12 ELISA-positive individuals out of 339 examined for *Leishmania* (namely, a seroprevalence of 3.6%; 95%-CI = 1.8%-5.7%), none showed active leishmaniasis clinical lesions. The presence of anti-*Leishmania* antibodies possibly reflects previous past exposure to the parasite or it indicates subclinical infections (26). It is noteworthy that most Venezuelan migrants passing through the Darién who attended the CMT of the UNAM, showed active lesions. All lesions were clinically suspicious for leishmaniasis, and several diagnostic tests were performed. Thus, from a sample of 26 migrants examined, 14 were positive for *Leishmania* (prevalence of 53%; 95%-CI = 35%-71%). Species identification was carried out by sequencing, which revealed that most patients were infected with *L.* (*V.*) *panamensis,* while only few cases involved *L.* (*V.*) *braziliensis*. Several species within the *Viannia* complex have been recorded in Panama, with *L.* (*V.*) *panamensis* being the dominant species (27).

*Lehismaniasis* (*V.*) *panamensis* is primarily distributed across Panama, Colombia, and Costa Rica. In contrast, *L.* (*V.*) *braziliensis* has a broader geographic range, predominating in South America (Brazil, Peru, Bolivia, Colombia, and Venezuela), although it has also been identified in parts of Central America (28).

*Nyssomyia* (*Nyssomyia*) *trapidoi*, *Ps. panamensis* and *Ny. ylephiletor* are the sand fly vectors associated with transmission of both *L.* (*V.*) *braziliensis* and *L.* (*V.*) *panamensis* (29–32). Currently, *Lu. gomezi* and *Lu. hartmanni* are associated with *L.* (*V.*) *panamensis* (10, 11), while *Pi. ovallesi* is associated with *L.* (*V.*) *braziliensis* (33). In Mexico, five vector species have been reported as capable of transmitting *L.* (*V.*) *braziliensis* (8).

Both *L.* (*V.*) *panamensis* and *L.* (*V.*) *braziliensis* are primarily associated with cutaneous leishmaniasis, characterized by chronic ulcerative lesions. However, *L.* (*V.*) *braziliensis* has also been strongly linked to mucocutaneous leishmaniasis (MCL), a rare but more severe clinical manifestation, with *L.* (*V.*) *panamensis* occasionally implicated as well (34–36).

The identification of these parasite species among migrant populations highlights the need for integrated epidemiological surveillance in both transit and destination areas, given the potential for introduction of pathogenic species in new regions. Furthermore, the clinical challenges to achieve a timely diagnosis and treatment remain a priority.

Taken together, these findings underscore the importance of integrating entomological and epidemiological surveillance within the migratory route analysis to advance further understanding of leishmaniasis dynamics in regions of South and Central America and Mexico. The convergence of competent vector species with susceptible vulnerable at-risk populations, such as migrants in transit, travelers, field workers, and military personnel, creates a complex epidemiological landscape and environment that requires a coordinated, multidisciplinary, and regionally informed epidemiological approach.

## Conclusions

The findings of this study confirm the presence of *L.* (*V.*) *panamensis* and *L.* (*V.*) *braziliensis* infections in migrants transiting through Mexico to the US from 2023 to 2024. Accurate species identification is crucial to help guide appropriate treatment and follow-up, reduce the risk of progression to mucocutaneous leishmaniasis (MCL), and advocate disease risks by supporting patient education on disease prevention and care. This study provides important evidence on the role of human migration in the epidemiology of leishmaniasis and underscores the need for an intersectoral integrated approach that incorporates social determinants of health alongside ecological and mobility-related factors. Integrating these dimensions is essential for the development of effective, sustainable, and regionally informed public health policies.

## Data Availability

All data will be available anytime to any scientist of institution upon request.

## Acknowledgments

We thank Itzel Shareny Espinosa-Ojeda for her valuable assistance in data cleaning of the migrant database. We also thank Miriam Anahi Andrade-Reyes (Instituto Politecnico Nacional), Yadira Gómez-Rodríguez, Julissa Domínguez-Osorio, and Nathalia Aimée Fernández-Santos for supporting field and office work.

